# Clinical characteristics of emergency patients with acute traumatic fractures

**DOI:** 10.1101/2025.04.10.25325178

**Authors:** Gengwei Zhang, Ling Zhang, Changming Xiao, Yuru Yang, Haifeng Chang

## Abstract

**Background:** Fractures are among the most common traumatic injuries worldwide, with distal radius fractures and femoral neck fractures being particularly prevalent[1,2]. These fractures not only affect patients’ quality of life but also impose a significant economic burden. Although there is extensive research on distal radius fractures and femoral neck fractures individually, comparative studies between the two are relatively scarce. Therefore, we analyzed clinical data from patients with distal radius fractures and femoral neck fractures treated at our hospital to explore the differences between these two types of fractures.

**Methods and findings:** This retrospective study included patients treated for distal radius fractures and femoral neck fractures at our hospital from January 2020 to February 2025. Data collected included gender, age, length of hospital stay, treatment methods, preoperative preparation time, and total hospitalization costs. Statistical analysis was performed on all data to analyze the characteristics of patients with distal radius fractures and femoral neck fractures.

During the study period, 204 patients with distal radius fractures and 124 patients with femoral neck fractures were included. Among distal radius fracture patients, 50% were male and 50% were female, with no statistically significant gender difference. Among femoral neck fracture patients, 45.16% were male and 54.84% were female, also with no statistically significant gender difference. The average age of distal radius fracture patients was 48.49 ± 14.51 years, while that of femoral neck fracture patients was 62.80 ± 17.64 years, showing a statistically significant difference. The average length of hospital stay for distal radius fracture patients was 8.31 ± 5.24 days, compared to 12.60 ± 6.96 days for femoral neck fracture patients, also statistically significant. Surgical treatment was performed in 80.88% of distal radius fracture patients and 80.65% of femoral neck fracture patients, with no statistically significant difference. The preoperative preparation time was 4.25 ± 2.55 days for distal radius fracture patients and 5.30 ± 3.31 days for femoral neck fracture patients, showing a statistically significant difference. The total hospitalization cost was 22,200.56 ± 13,295.27 yuan for distal radius fracture patients and 39,334.46 ± 27,283.70 yuan for femoral neck fracture patients, with a statistically significant difference.

**Conclusion:** This study revealed significant differences in clinical characteristics between patients with distal radius fractures and femoral neck fractures, particularly in age, length of hospital stay, preoperative preparation time, and hospitalization costs. Femoral neck fractures were more common in elderly patients, with longer preoperative preparation times, longer hospital stays, and higher treatment costs. In contrast, distal radius fracture patients were younger, with shorter preoperative preparation times, shorter hospital stays, and lower treatment costs.

## Introduction

Fractures are a common type of trauma in emergency departments worldwide. In clinical practice, distal radius fractures and femoral neck fractures are two important types of fractures. Distal radius fractures are the most common fractures in the upper limb, often resulting from falls where the hand is used to support the body. Femoral neck fractures are typically caused by osteoporosis or falls, and both types of fractures can lead to significant functional impairment. What are the differences in clinical characteristics between distal radius fractures and femoral neck fractures? By conducting an in-depth analysis of these differences, we can better understand the characteristics of patients with different types of fractures and provide a scientific basis for clinicians to develop appropriate treatment plans. Therefore, this paper will systematically compare these two types of fractures in terms of gender, age, length of hospital stay, treatment methods, preoperative preparation time, and total hospitalization costs, aiming to provide guidance for future clinical practice and research.

## Materials and Methods

### Study Subjects

This study was approved by the Ethics Committee of Shenzhen Third People’s Hospital. On February 25, 2025, following our request, the hospital provided patient data while protecting patient privacy and personal identity.This study was approved by the Ethics Committee of Shenzhen Third People’s Hospital. To ensure data accuracy and the reliability of the analysis results, we collected data from 204 patients with distal radius fractures and 124 patients with femoral neck fractures treated at our hospital from January 2020 to February 2025, while protecting patients’ personal information.

### Exclusion Criteria

Patients hospitalized multiple times for the same type of fracture were excluded; patients with incomplete information were excluded; patients with complex diseases (such as severe cardiovascular and pulmonary diseases, organ transplantation, etc.) were excluded; patients with old fractures were excluded; patients with open fractures were excluded; and patients with fractures in other parts of the body were excluded.

### Methods

Clinical data from the included cases were collected, including gender, age, admission date, discharge date, treatment methods, surgery date, and total hospitalization costs. A retrospective comparative analysis was conducted to compare the characteristics of patients with distal radius fractures and femoral neck fractures in terms of gender distribution, age, length of hospital stay, treatment methods, preoperative preparation time (time from admission to surgery), and total hospitalization costs.

### Statistical Analysis

Gender distribution and surgical treatment rates were categorical data, suitable for chi-square tests; for age, length of hospital stay, and preoperative preparation time, the data distribution did not meet the normality assumption, so non-parametric tests were used to determine differences; total cost data met the assumptions of independence, normality, and homogeneity of variance, so an independent samples t-test was used. A P-value <0.05 was considered statistically significant, and a P-value <0.01 was considered highly statistically significant.

## Results

### Gender Comparison

Among distal radius fracture patients, 102 were male (50.00%) and 102 were female (50.00%)(Fig1A), with a chi-square test P-value of 1.0. Among femoral neck fracture patients, 56 were male (45.16%) and 68 were female (54.84%)(Fig2A), with a chi-square test P-value of 1.0. This indicates no significant difference in gender distribution between distal radius fracture and femoral neck fracture patients (P>0.05;Table 1).

**Figure. 1.**
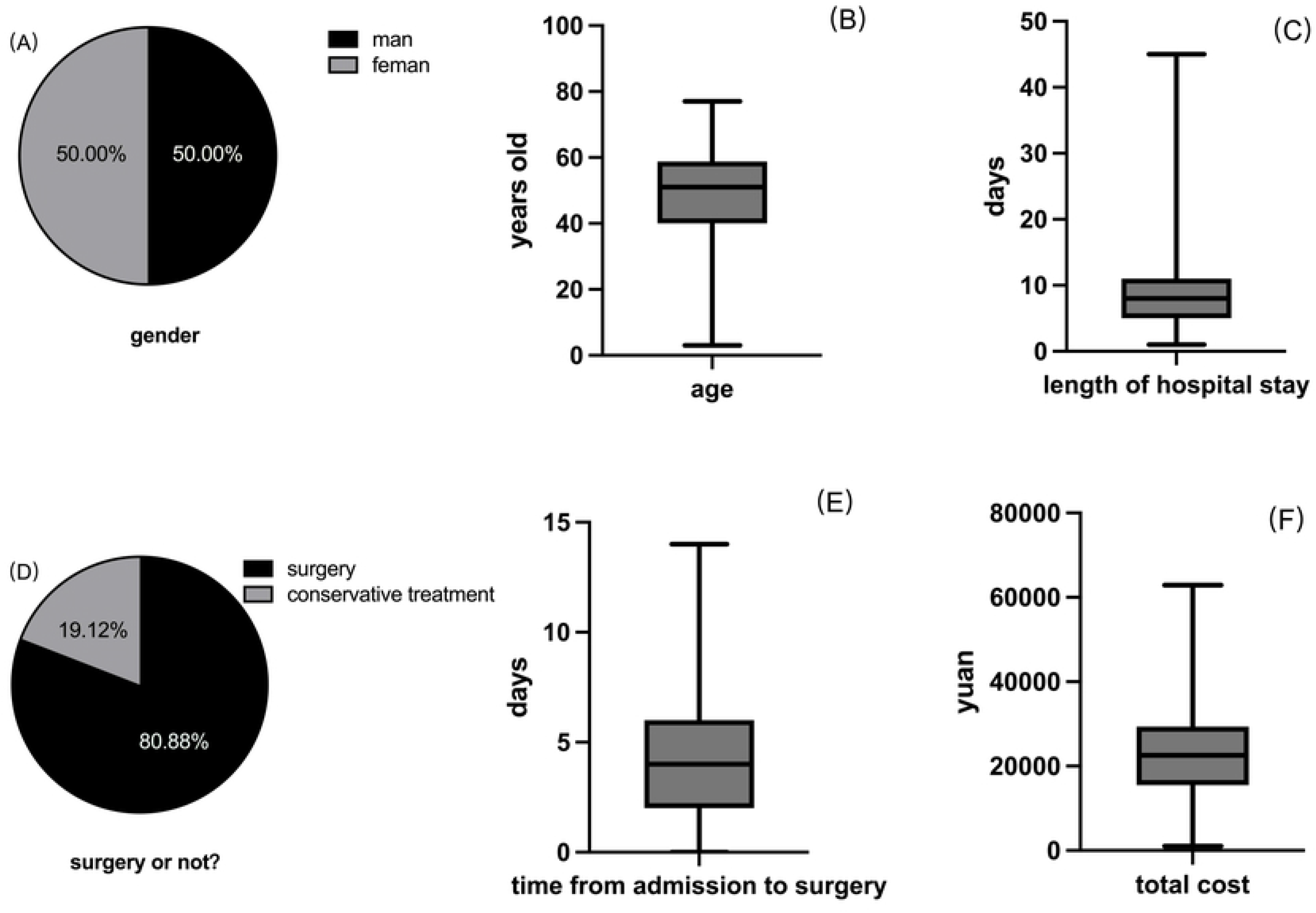
distal radius fractures

**Figure. 2.**
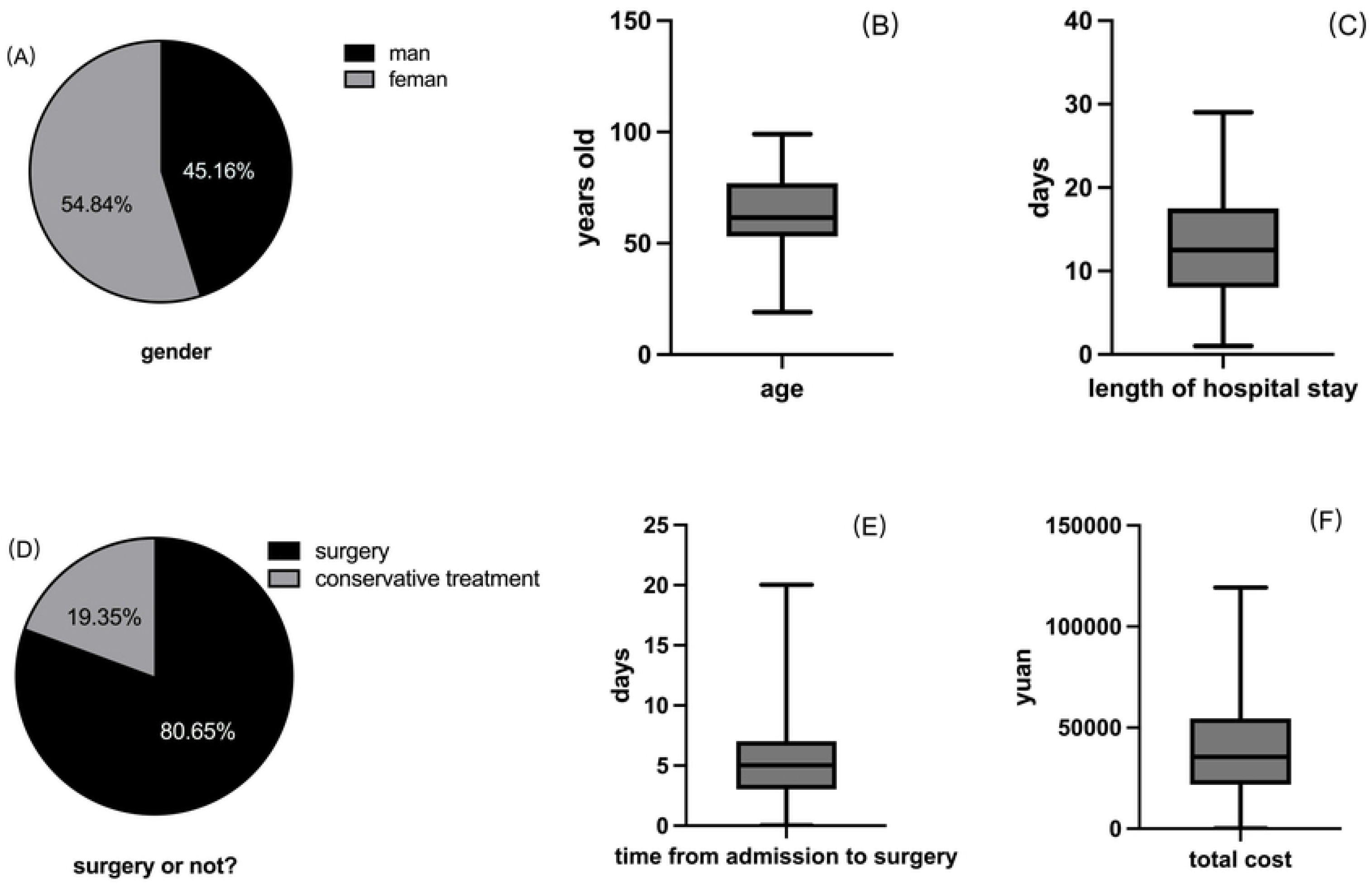
femoral neck fractures

**Table 1.** Gender Comparison.

| n (%) | male | female | Chi-square value | P-value |
| --- | --- | --- | --- | --- |
| distal radius fracture | 102(50.00) | 102(50.00) | 0.0 | 1.0 |
| femoral neck fracture | 56(45.16) | 68(54.84) | 0.0 | 1.0 |

### Comparison of Other Clinical Data

The average age of distal radius fracture patients was 48.49 ± 14.51 years(Fig1B), while that of femoral neck fracture patients was 62.80 ± 17.64 years(Fig2B). Based on normality test results, the data did not meet the normality assumption, so a Mann-Whitney U test was performed, showing a highly statistically significant difference in age between the two groups (P<0.001;Table 2), indicating that femoral neck fracture patients were older than distal radius fracture patients. The average length of hospital stay for distal radius fracture patients was 8.31 ± 5.24 days(Fig1C), compared to 12.60 ± 6.96 days(Fig2C) for femoral neck fracture patients. Based on normality test results, the data did not meet the normality assumption, so a non-parametric test (Wilcoxon rank-sum test) was performed, showing a highly statistically significant difference in length of hospital stay between the two groups (P<0.001;Table 2), indicating that femoral neck fracture patients had longer hospital stays than distal radius fracture patients. Surgical treatment was performed in 165 distal radius fracture patients (80.88%)(Fig1D) and 100 femoral neck fracture patients (80.65%)(Fig2D), with a chi-square test P-value >0.05, indicating no significant difference in surgical treatment rates between the two groups (Table 2). The preoperative preparation time was 4.25 ± 2.55 days(Fig1E) for distal radius fracture patients and 5.30 ± 3.31 days(Fig2E) for femoral neck fracture patients. Based on normality test results, the data did not meet the normality assumption, so a Mann-Whitney U test was performed, showing a statistically significant difference in preoperative preparation time between the two groups (P=0.006;Table 2), indicating that femoral neck fracture patients had longer preoperative preparation times. The total hospitalization cost was 22,200.56 ± 13,295.27 yuan(Fig1F) for distal radius fracture patients and 39,334.46 ± 27,283.70 yuan(Fig2F) for femoral neck fracture patients. The cost distribution for femoral neck fracture patients was more dispersed, and an independent samples t-test showed a highly statistically significant difference in total costs between the two groups (P<0.001;Table 2), indicating that femoral neck fracture patients had significantly higher treatment costs than distal radius fracture patients.

**Table 2.**
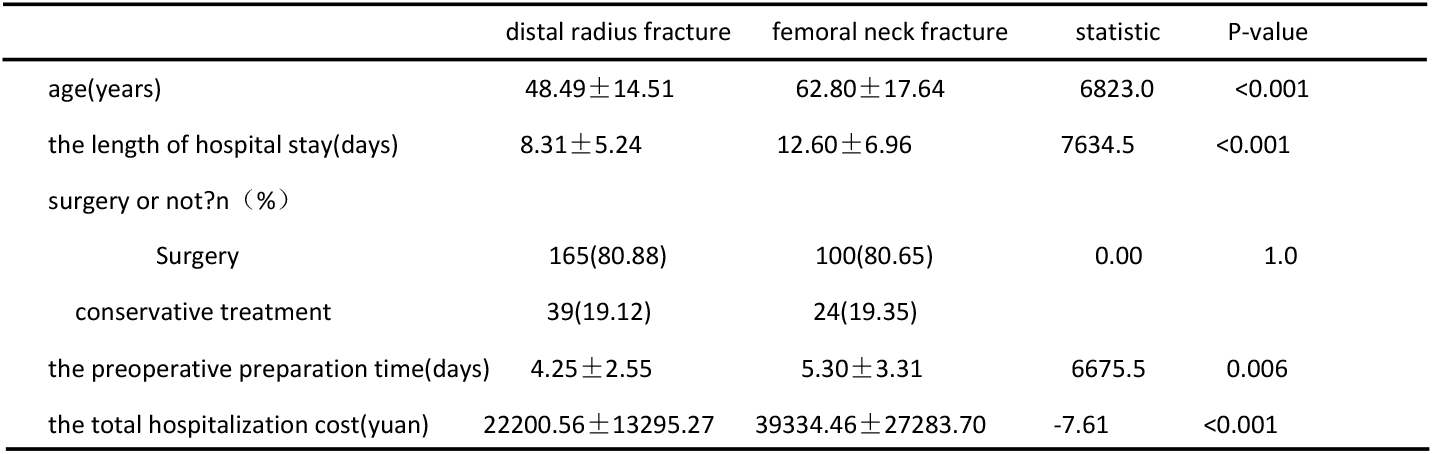
Comparison of Other Clinical Data.

|  | distal radius fracture | femoral neck fracture | statistic | P-value |
| --- | --- | --- | --- | --- |
| age(years) | $48.49 \pm 14.51$ | $62.80 \pm 17.64$ | 6823.0 | <0.001 |
| the length of hospital stay(days) | $8.31 \pm 5.24$ | $12.60 \pm 6.96$ | 7634.5 | <0.001 |
| surgery or not?n (%) |  |  |  |  |
| Surgery | 165(80.88) | 100(80.65) | 0.00 | 1.0 |
| conservative treatment | 39(19.12) | 24(19.35) |  |  |
| the preoperative preparation time(days) | $4.25 \pm 2.55$ | $5.30 \pm 3.31$ | 6675.5 | 0.006 |
| the total hospitalization cost(yuan) | $22200.56 \pm 13295.27$ | $39334.46 \pm 27283.70$ | -7.61 | <0.001 |

## Discussion

This study, through a comparative analysis of clinical characteristics in patients with distal radius fractures and femoral neck fractures, identified significant differences in age, length of hospital stay, preoperative preparation time, and total hospitalization costs, providing important insights for clinical practice and further research.

The results of this study showed no significant difference in gender distribution between femoral neck fracture and distal radius fracture patients. According to the literature[3], the proportion of female patients with femoral neck fractures is typically significantly higher than that of male patients, mainly due to the higher incidence of osteoporosis in elderly women. In contrast, the gender distribution of distal radius fractures remains controversial, with some studies showing a higher proportion of female patients[4], while others indicate no significant gender difference[5], with this study supporting the latter.

The results of this study showed that the average age of femoral neck fracture patients was significantly higher than that of distal radius fracture patients, consistent with most previous studies[6,7]. Femoral neck fractures are often caused by low-energy injuries, and elderly individuals are more prone to falls due to decreased balance, muscle strength[8], vision, and cognitive function. In contrast, distal radius fractures can occur in all age groups, with younger patients often experiencing high-energy injuries (such as sports injuries or traffic accidents)[9]. This difference in injury mechanisms directly leads to significant differences in age distribution between the two types of fractures. Additionally, our hospital rarely treats pediatric fractures, and if more pediatric cases were included, the age difference between the two types of fractures might be further amplified.

In terms of length of hospital stay, this study showed that femoral neck fracture patients generally had longer hospital stays. This phenomenon is mainly due to the complexity of femoral neck fracture surgery and the longer postoperative rehabilitation. In contrast, the treatment of distal radius fractures is relatively simple, resulting in shorter hospital stays. In clinical practice, appropriate rehabilitation plans should be developed for different fracture patients to ensure optimal treatment and care during hospitalization.

This study found no significant difference in the proportion of surgical treatment between patients with femoral neck fractures and distal radius fractures, which is inconsistent with previous studies[10, 11]. This discrepancy is likely due to the fact that this study only included data from hospitalized patients. Patients with femoral neck fractures are often hospitalized, whereas the hospitalization rate for distal radius fractures is relatively low[12]. Non-hospitalized patients with distal radius fractures typically have stable fractures, for whom cast immobilization is the preferred treatment option, and they can complete follow-up on an outpatient basis. Therefore, this study did not encompass data from these non-hospitalized patients. Future research should expand the sample scope to include outpatient data, thereby providing a more comprehensive reflection of the current treatment landscape for both types of fractures.

Femoral neck fracture patients had significantly longer preoperative preparation times than distal radius fracture patients, which may be related to patient age, as the proportion of elderly patients in the femoral neck fracture group was significantly higher than in the distal radius fracture group. Elderly patients often have multiple comorbidities, such as cardiovascular disease, diabetes, and respiratory diseases[13], making preoperative assessment more complex. Femoral neck fracture patients require more specialist consultations and auxiliary examinations, directly leading to longer preoperative preparation times. In particular, cardiac function assessment, pulmonary function testing, and nutritional status evaluation are essential for elderly patients, along with medication adjustment and anesthesia risk assessment. These factors are likely the main reasons for the longer preoperative preparation times in femoral neck fracture patients.

Finally, the results of this study showed that the total hospitalization costs for femoral neck fracture patients were significantly higher than those for distal radius fracture patients, consistent with previous studies[14,15,16]. We believe this difference is mainly due to the combined effects of preoperative preparation, surgical costs, and length of hospital stay. Femoral neck fracture patients are predominantly elderly, and their preoperative preparation includes multiple examinations such as cardiac function, pulmonary function, and nutritional status. These additional examinations directly increase medical costs. Femoral neck fracture surgery often involves internal fixation or joint replacement, which is more complex and involves expensive materials. The length of hospital stay for femoral neck fracture patients is significantly longer than that for distal radius fracture patients, and the extended hospital stay not only increases bed costs but may also lead to more complications and subsequent treatment costs. In clinical practice, doctors should develop individualized treatment plans based on patients’ specific conditions, optimizing resource allocation and reducing medical costs while ensuring the quality of care.

## Data Availability

This study is a review article and does not contain any original research data.

## Supporting information

Original data.

## Acknowledgments

This study was supported by the Shenzhen Clinical Medical Research Center for Trauma Treatment Fund [LCYSSQ20220823091405012].

## Author Contributions

Data curation: Ling Zhang.

Formal analysis: Changming Xiao.

Investigation: Yuru Yang.

Writing–original draft: Gengwei Zhang.

Writing–review and editing: Haifeng Chang.

